# Factors contributing to immunization coverage among children less than 5 years in Nadowli-Kaleo District of Upper West Region, Ghana

**DOI:** 10.1101/2024.01.17.24301424

**Authors:** Alice Kuuyi, Robert Kogi

## Abstract

Immunization is believed to prevent deaths from diseases such as diphtheria, tetanus, whooping cough and measles in about 2.5 million children each year worldwide. Failure to vaccinate children in the required timeframe could result in disease outbreaks and increased costs. This study was to identify the causes of low vaccination coverage among children in Nadowli-Kaleo district, Ghana.

An analytical cross-sectional study was adopted for this study. Three hundred and thirty-six participants (336) were recruited through a systematic sampling method. Data was collected using KoboCollect online data collection tool. Data was analyzed using Stata Version 17.0. Chi-square test was used to establish a link between the child’s immunization status and the independent variables. Finally, logistic regression was used to determine the degree of association. To determine statistical significance, a p-value of less than 0.05 at the 95 percent confidence level was employed.

It was revealed that immunization status among children under-five was a little above average (55.4%). Factors that were significantly associated with child full immunization were mother’s or caregiver’s age, marital status, occupation, and current child’s birth order (p-value < 0.05). Other location-related factors significantly associated with full immunization of children were number of ANC visits, delivery location, and distance to health facility. Giving birth to a child at the health facility had higher chance of 2.12 times to be fully immunized than giving birth at home [AOR (95%CI) p-value=2.12 (1.14-3.94)0.017]. The health system related factors which were found to be statistically associated with child full immunization included time spent during immunization service and being informed when to come for the next vaccination.

Immunization coverage is not very encouraging at our study site. To enhance childhood vaccination rates, we suggest expanding access to health services, institutional childbirth, and timely regular antenatal visits.

## Introduction

Immunization is a measure used in a person to make him or her resistant against an infectious disease by typically administrating a vaccine or previous infection to the wild microorganism [1]. Vaccine is given to protect the person’s body system against any future infection or disease [2]. Immunization coverage, on the other hand, is referred to the percentage of individuals in a target population who are vaccinated [3]. Over the years, immunization has been proven to be a very cost-effective and life-saving intervention which protects people from needless suffering like sickness, disability, and death. Most importantly, the benefits of immunization are to all people, which target the improvement in health and life expectancy, with impact at the social, economic global, national, and community levels [4]. There is a great contribution to be made by the increasingly interdependent communities working together to tackle diseases, which are of public health concern and can be prevented with vaccine [5].

Basic immunizations are estimated to avert 2.5 million annual child deaths globally from diphtheria, tetanus, Pertussis and measles [6]. Currently, more children are getting vaccinated at the appropriate time, but approximately 20 million individuals across the globe are still not receiving vaccinations, leaving them vulnerable to severe illnesses, fatalities, handicaps, and poor health. African regions alone accounted for 8.5% of unvaccinated children in the world and almost much as in other regions combined [7]. Globally, Africa has the lowest coverage of children fully vaccinated [8]. A weakness of this system is the high dropout rate between doses. Therefore, there is a need to strengthen health care and routine immunization systems throughout the region [9].

It was established in Ghana in 2014 that, nationwide, the proportion of children aged 12-23 months who did not receive all basic immunizations decreased from 79% to 77% [9]. During the same period, the proportion of children without childhood vaccination increased nationally from 1% to 2% [9].

The need to raise child survival is a key global development goal, which is emphasized once again in the Sustainable Development Goals (2016–2030). Massive investments have been made as a result of these agreements to enhance the acceptability, affordability, and accessibility of child health intervention programs, particularly those related to nutrition and immunization services [10]. The Ghana Health Service (GHS) has enhanced its Expanded Programme on Immunization (EPI) to align with the global objectives established at the 1988 World Health Assembly. In addition to vaccination coverage, the GHS now emphasizes eradicating other illnesses like poliomyelitis, while also striving to eliminate measles and neonatal tetanus. Disease surveillance and control measures have also been intensified [11]. Immunization performance has become a key health performance indicator in Ghana for the entire health sector and is monitored at all levels [11]. However, reports show that some districts record low immunization, stifling the achievement of set targets.

Studies have indicated that factors that affect immunization coverage could be client-related, service-related, or health management-related. For instance, it was reported that lack of information or motivation, place of immunization not convenient for mothers, inconvenient immunization time, vaccinators absent, family problems, including maternal overwork, maternal illness and child illness, were identified as factors which influenced immunization coverage [12]. The level of compliance of mothers with childhood immunization is also influenced by their knowledge of childhood immunization, higher level of maternal education, birth within close proximity to a medical facility and in a medical facility [13].

The national target for all antigens in Ghana is 95% coverage [14]. Penta 3 has been used as a reference indicator for measuring the coverage, quality, and utilization of childhood immunization. For the past 10 years, Ghana is able to maintain high immunization coverage rates between roughly 90-95%. However, there are reports of low-performing areas in some districts which are not achieving the 95% target with 3 doses of pentavalent vaccine [15]. This has been demonstrated in the Nadowli-Kaleo district where the immunization coverage has been consistently below the national target of 95% coverage for all antigens over the five years. The Penta 3 coverage for Nadowli-Kaleo District from 2015 to 2019 was 71.9%, 71.3%, 70%, 70% and 72.7% respectively [16]. This poses a threat to vaccine-preventable disease outbreaks and other implications of low coverage, hence, making this study necessary.

Enhancing the extent of immunization is crucial in advancing the health of children and diminishing the incidence of ailments and fatalities during childhood. However, immunization coverage in developing countries has been reported to be low [6]. For instance, in Ethiopia, a study conducted using a cross-sectional study design revealed that 77.4% of children received full immunization, 15.5% children were partially immunized, while some 7.1% did not receive any antigen (6). In Mukurweni and Tetu Sub-counties, Kenya, moderate and high immunization coverages were reported in Mukurweini and Tetu Divisions respectively [17].

Moreover, it has been found that substantial socioeconomic inequities, such as residency, wealth, educational status, and the number of children in a household, affect the coverage of immunization [18]. For instance, in Kenya, a population-based cross-sectional study conducted identified that the factors which influenced low immunization coverage included; family size, location of child’s birth, the level of literacy, awareness of the vaccination schedule, and whether or not the family lives a nomadic lifestyle [5]. Moreover, Kiptoo and colleagues noted that the income of the household, the proximity of the nearest healthcare facility, and whether the family resided in an urban or rural area impacted immunization coverage [5].

In addition, another study conducted in the slums of Mumbai, India, using a community-based cross-sectional approach found that inadequate time, insufficient knowledge, concerns about negative side effects, and potential income loss were among the significant factors that hindered people from receiving immunizations [19]. Moreover, a study conducted in Burkina Faso revealed that factors such as location of residence, year of visit, ethnic group, and maternal education were strongly linked to the vaccination status of individuals [20]. Additionally, factors related to the mother such as her occupation, educational background, age, and familiarity with vaccine-preventable illnesses and vaccination, were identified as influencers of immunization coverage in the Assin North Municipality, located in the Central Region of Ghana [21]. Also, it was revealed in another study conducted in the Tamale Metropolis that attaining a higher education, being a mother, having an older age, earning a higher income, having a greater number of children, living in rural areas, possessing extensive knowledge on immunization, and exhibiting a positive attitude towards immunization, all resulted in a decline in the failure to comply with all the planned immunizations [22].

It was further reported in India that caregivers perceived several barriers to immunization. These barriers included a shortage of time, concerns about potential adverse effects from the vaccination, and poor staff conduct [19]. In Ethiopia, it was also noted that the distance from health facility, delivery place, and follow-up during ANC, were significance with a child being fully immunized [23]. Similarly, it was revealed in a qualitative study conducted in Ethiopia that the most commonly shared factors among the discussants which affected routine childhood immunization service uptake were inaccessible health facilities, unfavourable attitudes, poor performance and mistreatment of healthcare personnel, inconvenient vaccination schedules, insufficient information during vaccination days, and extended waiting periods [23].

## Methods

### Study Design

This study adopted a cross-sectional study involving 336 mothers whose children were under five years of age and living in the Nadowli-Kaleo District. This design was chosen as the researchers intended to gain knowledge and information on low immunization coverage within a short period in the study area.

### Study Site Description

The Nadowli-Kaleo District is positioned in the central region of Ghana’s Upper West area, with latitude between 10’ 20’ and 11’ 30’ north and longitude 3’10’ and 2’10’ west. It shares borders to the south with the Wa Municipal, to the west with Burkina Faso, north with Jirapa District, and to the east with the Daffiama-Bussie-Issa District. The district spans an area of 2,742.50 km2, stretching from the Billi Bridge to the Dapuori Bridge, nearly 12 km away from Jirapa. The district extends from the Black Volta (Charikpong) to Daffiama from west to east. According to the 2010 Population and Housing Census, the district had a population of 61,561, which accounted for 8.8% of the Upper West Region’s total population (24). The majority (53.35%) of the population of the district is the female group with a Total Fertility Rate (TFR) of 3.2 [24]. On the other hand, the General Fertility Rate (GFR) stood at 85.8 births per 1000 women in fertility age (15-49 years), which is the fourth lowest among the districts in the region [24]. Finally, the Nadowli-Kaleo district has one district hospital serving as a referral centre situated in Nadowli, 10 health centres, and 145 outreach points which offer immunization services.

### Study Population

All mothers or caregivers with children below the age of five and living within the Nadowli-Kaleo District were part of the study’s participants.

### Inclusion and Exclusion criteria

The mothers or caregivers who were included in this study were those with children up to five years of age, who were residents of the district. Mothers whose children fit the inclusion criteria but were severely sick and needed medical attention were excluded.

### Sample Size Determination

We calculated the sample size for this study by using the Cochran formula (25), 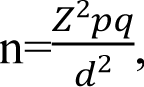 Where n= Sample size, Z = Z_score_, p= proportion estimated for immunization coverage present in the population. Coverage of 72.7% obtained in the year 2019 for Penta 3 [14] in the Nadowli-Kaleo District is used. q = 1-p, d= Level of statistical significance of 95% = (0.05). The used the assumption of a margin of error of 0.05 and a 95% confidence level.

A sample size 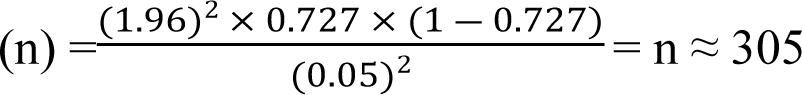

Adding a non-respond rate of 10%, n= (305*0.10) +305 = 336. Therefore, 336 mothers with children less than five years were considered in the study.

### Sampling method

All eight subdistricts in the Nadowli-Kaleo District were considered in conducting this study. This study was conducted in health centres because they recorded a high intake of CWC attendance in the district. The data was collected in all 8 subdistricts in the district, and all 9 health centres where CWC was conducted were purposively used.

The proportionate sample size at each CWC centre was obtained using the 2019 immunization coverage for Penta 3 per subdistrict as shown in Table 1. These proportional samples were calculated and estimated in the table below.

**Table 1:** Proportional allocation of study population by Sub-district

| Sub-district | Immunization Coverage | Sub-district sample size = $\frac{\text{Sub - district coverage}}{\text{District coverage}} \times \text{study sample size}$ | |
| --- | --- | --- | --- |
| Charikpong | 210 | $\frac{210}{2166} \times 336 = 33$ | |
| Dapour | 186 | $\frac{186}{2166} \times 336 = 29$ | |
| Jang | 406 | $\frac{406}{2166} \times 336 = 63$ | |
| Kaleo | 415 | $\frac{415}{2166} \times 336 = 64$ | |
| Nadowli | 409 | $\frac{409}{2166} \times 336 = 63$ | |
| Nanvil | 120 | $\frac{120}{2166} \times 336 = 19$ | |
| Sombo | 187 | $\frac{187}{2166} \times 336 = 29$ | |
| Takpo | 233 | $\frac{233}{2166} \times 336 = 36$ | |
| **Total** | **2166** | **336** |  |

To obtain the appropriate sample size for this study, a systematic sampling technique was used. Systematic sampling is a type of random sampling where the initial unit is chosen randomly using random numbers, and subsequent units are chosen automatically based on a predetermined pattern (26). Initially, the researchers collaborated with the heads of different healthcare facilities to determine the count of children registered in the Child Welfare Clinic (CWC) registers who fell within the age range of 0 to 59 months. The register was numbered serially to form the sampling frame, then the proportionate sample to be obtained was used to divide it. Secondly, the total number of children in each register was divided by the respective samples for each of the subdistricts to calculate the interval that was used when selecting the children. Numbers on the ballot papers were given from 1 to the interval number that was obtained. That number corresponded to the mother who had that number that comes with the facility. That sampling interval number was then be added to obtain every nth number.

### Data collection procedure

Four people were trained to collect the data. The training was done using both English and Dagaare; demonstration was done using Dagaare which aided in effective data collection. The essence of the research and procedure were explained to them. They were encouraged to ask questions to clarify any doubts.

Interviewer-administered pretested questionnaire was administered to respondents using Kobo Collect online data collection tool. The questionnaire was made up of five sections: socio-demographic characteristics of respondents, knowledge-based questions of caregivers/mothers on child immunization, and attitude of caregivers/mothers towards child immunization. To obtain health system related factors, mothers were asked such questions as long waiting period, attitude of health workers towards them, special education to them on immunization when they came, information on side effects, how to take care of it, as well as distance to the nearest facility. Data was collected on the days’ caregivers/mothers attended the child welfare clinic. The methods employed to distribute the questionnaires involved individualized interaction, where the questions were asked directly and answered by the respondents. On average, it took approximately 15 minutes for each respondent to complete the questionnaire.

### Data analysis

The data produced was processed and imported into the STATA 17.0 software for analysis. The main focus of the study was to determine the immunization status of children under the age of 5, which was classified as either fully immunized or partially immunized. In addition, the researchers analyzed independent variables, including factors related to the mother’s individual and community characteristics such as age, education, occupation, antenatal care visits, and the birth order of the child. Categorical variables were presented as proportions and frequencies. The dependent/outcome variable was a dichotomous variable (Fully or partially immunized). Immunization was defined as follows: 1 is full immunization if the child received both doses of the vaccine as recommended and partially as 0 if a child did not receive any dose or received one dose of vaccine before the study. The association between immunization status and independent variables was analyzed using the chi-square test. Additionally, logistic regression was employed to assess the predictors of the immunization. To determine significant associations, a confidence interval of 95% and a p-value below 0.05 were used as criteria.

### Ethical Issues

Ethical clearance was sought from University of Health and Allied Sciences (UHAS) Ethics Review Committee for approval (**UHAS-REC A.9** [126] **20-21**). Permission was sought from the Upper West Regional Directorate and the District Health Directorate of the Nadowli-Kaleo District. Formal verbal consent was obtained from all the participants involved in the study. The recruited participants were given complete information regarding the research’s aim, and their privacy as well as confidentiality were guaranteed. The researchers sought the consent from all the participants. It is worth noting that no mother or caretaker was compensated for their involvement in the study. However, they were assured that the findings from this study would be used to improve the overall immunization coverage in the district.

Any interaction during data collection was done while wearing a facemask and maintaining social distancing as ways of observing the Covid-19 protocol. Research assistants and participants were also used alcohol-based hand sanitizers throughout the period of the data collection.

## Results, Discussion, Conclusions

### Results

The results in Table 2 showed that the majority, 144 (42.9%) of the respondents (mothers) were 30 years old and below. This was immediately followed by those aged 31-35 years, 141 (42.0%). A greater proportion, 193 (57.4%) of the children in this study were within 24-59 months old and most, 173 (51.5%) were females. Majority 147 (43.8%) of the children in this study were first born.

**Table 2:**
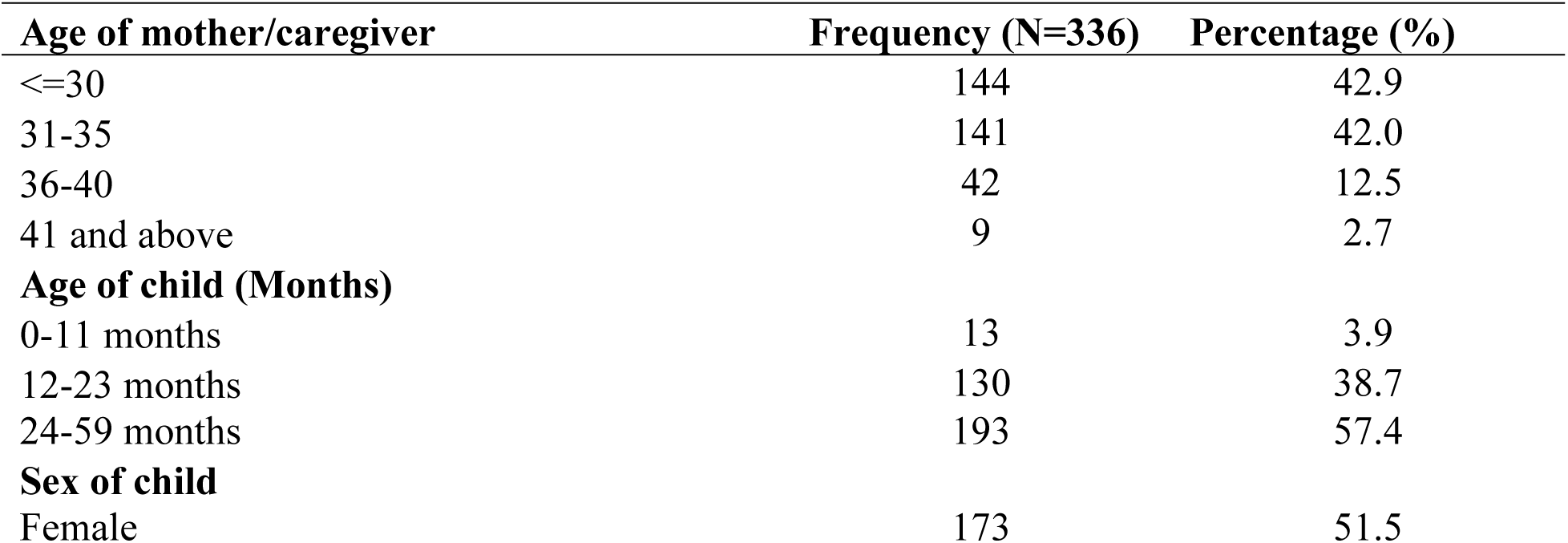
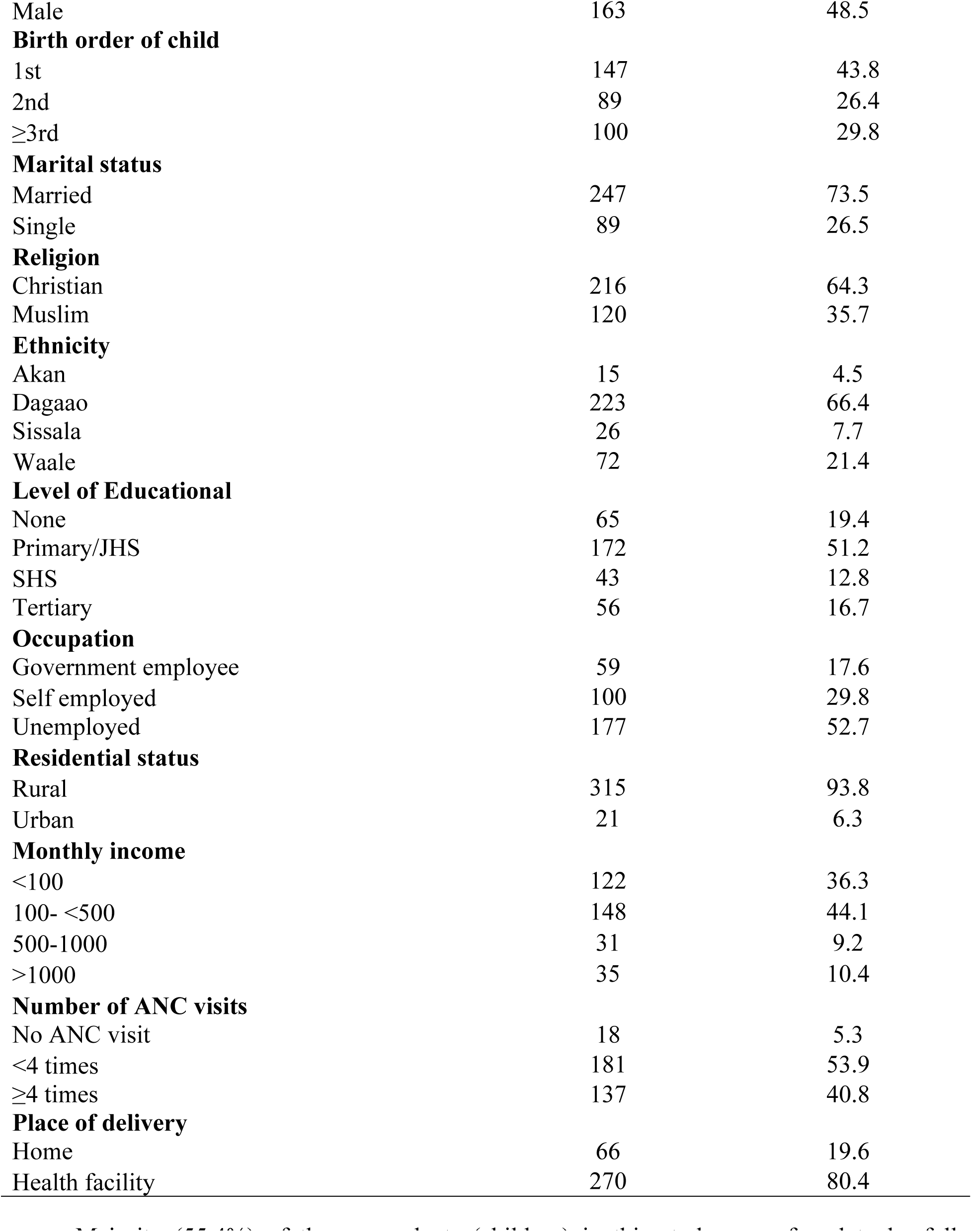
Socio-demographic characteristics of mothers/caregivers

Among the mothers, a majority 247 (73.5%) of them were married and a greater number 216 (64.3%) of the mothers were Christians. Moreover, most 223 (66.4%) of the respondents were Dagaabas. Majority 172 (51.2%) of the respondents attained primary or JHS education. The results further showed that majority 177 (52.7%) of the respondents were unemployed and most 315 (93.8%) of them were rural dwellers. In addition, majority 148 (44.1%) of the respondents earned from Gh₵100.00-<Gh₵500.00. Moreover, the majority 181 (53.9%) of the mothers in this study were found to make four ANC visits. Finally, a vast majority 270 (80.4%) of the mothers delivered in a health facility.

Majority (55.4%) of the respondents (children) in this study were found to be fully immunized as indicated in Fig 1.

**Fig 1:**
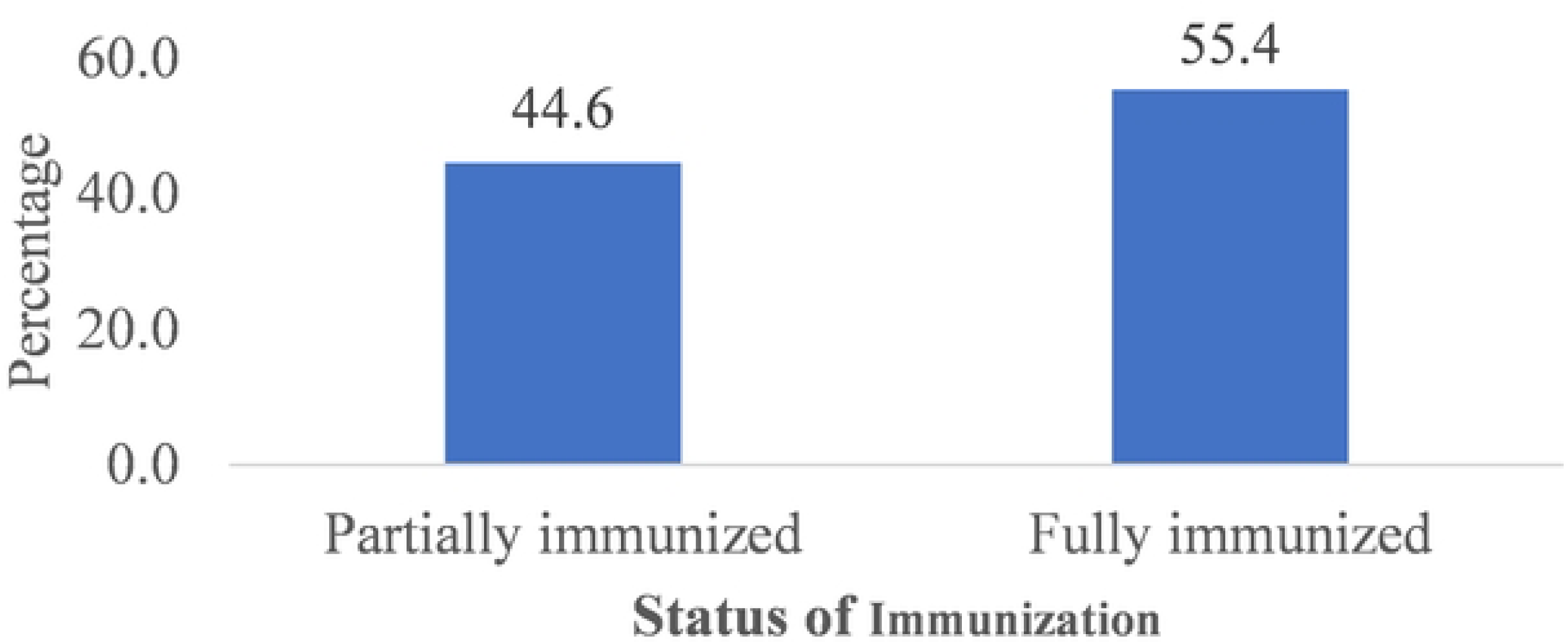
Immunization status of children

It is found in Table 3 that the majority 85 (45.7%) of the respondents who were fully immunized were within 31-35 years and most 149 (80.1%) of the women who were married had their children fully immunized. Moreover, most 116 (62.4%) Christians had their children fully immunized. A greater proportion 122 (65.6%) of the respondents who belonged to the Dagaaba ethnic group had their children fully immunized. A higher proportion 178 (95.7%) of the respondents who were rural dwellers had their children fully immunized.

**Table 3:** Association between respondents’ socio-demographic characteristics and child full immunization

| Variables | Immunization status |  | Pearson Chi-square | p-value |
| --- | --- | --- | --- | --- |
|  | Partially immunized<br>(n=146) | Fully immunized<br>(n=190) |  |  |
| **Age (Mother/caregiver)** |  |  |  |  |
| ≤30 | 77 (51.3) | 67 (36.0) | 8.57 | 0.014 |
| 31-35 | 56 (37.3) | 85 (45.7) |  |  |
| 36 and above | 17 (11.3) | 34 (18.3) |  |  |
| **Marital status** |  |  |  |  |
| Married | 98 (65.3) | 149 (80.1) | 9.31 | 0.002 |
| Single | 52 (34.7) | 37 (19.9) |  |  |
| **Religion** |  |  |  |  |
| Christian | 100 (66.7) | 116 (62.4) | 0.67 | 0.413 |
| Muslim | 50 (33.3) | 70 (37.6) |  |  |
| **Ethnicity** |  |  |  |  |
| Dagaao | 101 (67.3) | 122 (65.6) | 1.31 | 0.520 |
| Sissala | 15 (10.0) | 26 (14.0) |  |  |
| Waale | 34 (22.7) | 38 (20.4) |  |  |
| **Level of Educational** |  |  |  |  |
| None | 26 (17.3) | 39 (21.0) | 6.92 | 0.075 |
| Primary/JHS | 82 (54.7) | 90 (48.4) |  |  |
| SHS | 24 (16.0) | 19 (10.2) |  |  |
| Tertiary | 18 (12.0) | 38 (20.4) |  |  |
| **Occupation** |  |  |  |  |
| Government employee | 17 (11.3) | 42 (22.6) | 7.82 | 0.020 |
| Self employed | 51 (34.0) | 49 (26.3) |  |  |
| Unemployed | 82 (54.7) | 95 (51.1) |  |  |
| **Residential status** |  |  |  |  |
| Rural | 137 (91.3) | 178 (95.7) | 2.70 | 0.100 |
| Urban | 13 (8.7) | 8 (4.3) |  |  |
| **Monthly income** |  |  |  |  |
| <500 | 140 (93.3) | 165 (88.7) | 2.12 | 0.145 |
| ≥500 | 10 (6.7) | 21 (11.3) |  |  |
| **Age of child (Months)** |  |  |  |  |
| ≤12 | 11 (7.5) | 10 (5.3) | 0.73 | 0.394 |
| >12 | 135 (92.5) | 180 (94.7) |  |  |
| **Sex of children** |  |  |  |  |
| Female | 75 (51.4) | 98 (51.6) | 0.00 | 0.970 |
| Male | 71 (48.6) | 92 (48.4) |  |  |
| **Birth order of current child** |  |  |  |  |
| 1st | 74 (49.3) | 73 (39.3) | 7.90 | 0.019 |
| 2nd | 43 (28.7) | 46 (24.7) |  |  |
| ≥3rd | 33 (22.0) | 67 (36.0) |  |  |

Moreover, the majority 90 (48.4%) of the respondents who attained either primary or JHS education had their children fully immunized. Majority 95 (51.1%) of the unemployed were found to have their children fully immunized and most 165 (88.7%) of the respondents who earned less than GH₵500.00 a month had their children fully immunized. Moreover, most 180 (94.7%) of the children who were above 12 months of age were fully immunized. Additionally, the majority 98 (51.6%) of the children who were females were fully immunized, while most 73 (39.3%) of the first-born children were fully immunized.

Furthermore, it was found in this study that the factors which were significantly associated with child’s full immunization were mother’s or caregiver’s age, marital status, occupation, and birth order of the current child [X^2^(P-value =8.57(0.014), 9.31(0.002), 7.82(0.020), and 7.90(0.019) respectively].

The results in Table 4 showed that the majority 90 (47.4%) of the respondents who made at least one ANC attendance had their children fully immunized. For respondents who delivered in the health facility, majority, 166 (87.4%) of them had their children fully immunized. Mothers who walked at most 5 kilometres to the nearest health facility had most of their children being fully immunized 169 (88.9%). Additionally, the results further depicted that number of ANC visits, place of delivery, and distance to health facility were all factors which had statistically significance association with child full immunization [X^2^(P-value) =10.13(0.006), 13.62(P<0.001) and 4.38(0.036) respectively].

**Table 4:** Association between children’s immunization status and community-related factors.

| Immunization status |  |  |  |  |
| --- | --- | --- | --- | --- |
| Variables | Partially immunized<br>(n=146) | Fully immunized<br>(n=190) | Pearson Chi-square | p-value |
| Number of ANC visits |  |  |  |  |
| No ANC visit | 11 (7.5) | 10 (5.2) | 10.13 | 0.006 |
| ≤3 times | 89 (61.0) | 90 (47.4) |  |  |
| 4+ times | 46 (31.5) | 90 (47.4) |  |  |
| Place of delivery |  |  |  |  |
| Home | 42 (28.8) | 24 (12.6) | 13.62 | P<0.001 |
| Health facility | 104 (71.2) | 166 (87.4) |  |  |
| Distance to health facility |  |  |  |  |
| >5KM | 28 (19.2) | 21 (11.1) | 4.38 | 0.036 |
| ≤ 5KM | 118 (80.8) | 169 (88.9) |  |  |

The results in Table 5 showed that the majority 99 (52.1%) of the respondents who indicated that they were never screamed at by health worker (s) when they were late during immunization had their children fully immunized. Moreover, most 115 (60.5%) of the mothers who spent less than 30 minutes in immunization had their children fully immunized. A greater proportion, 146 (76.8%) of the mothers who were educated on how to manage vaccine side effects also had their children fully immunized. Lastly, the majority 153 (80.5%) of the mothers who were informed about their next CWC visit also had their children fully immunized.

**Table 5:**
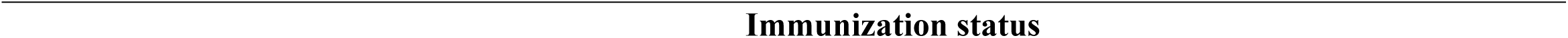
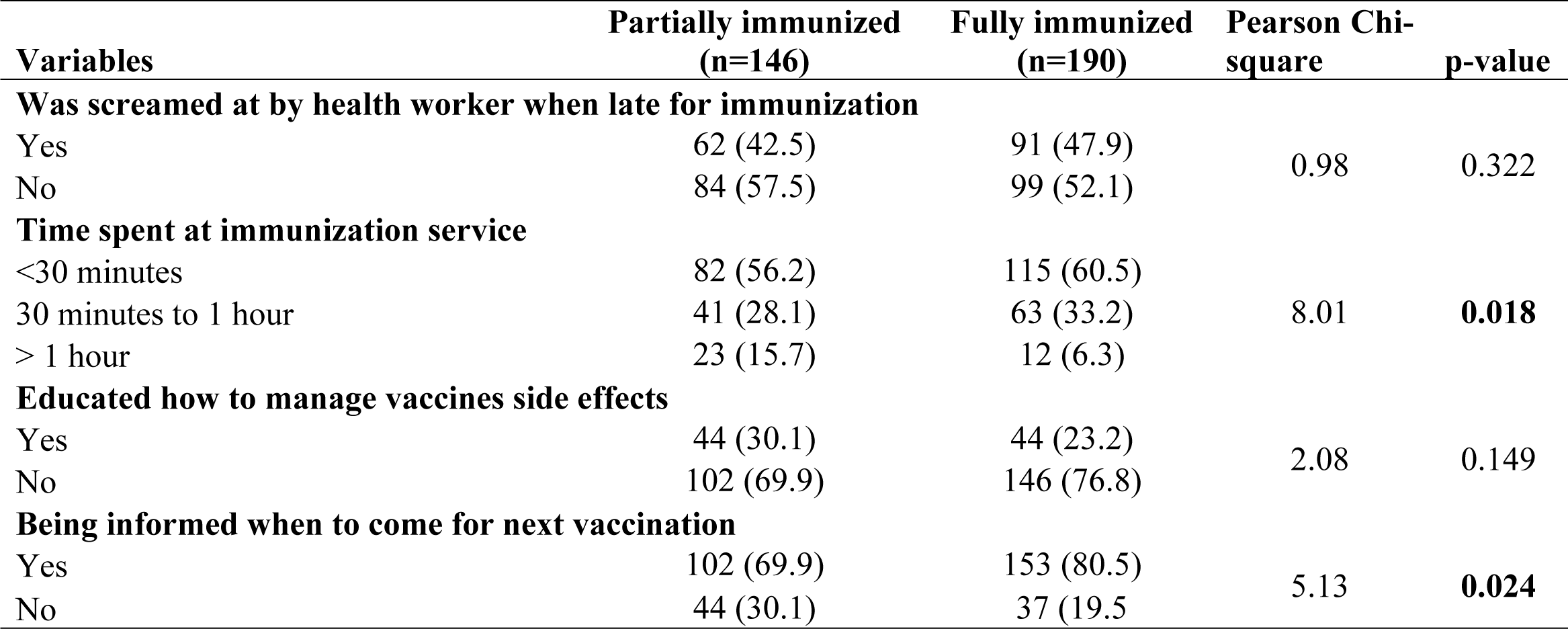
Association between child immunization status and health system-related factors

Immunization status
| Variables | Partially immunized<br>(n=146) | Fully immunized<br>(n=190) | Pearson Chi-square | p-value |
| --- | --- | --- | --- | --- |
| **Was screamed at by health worker when late for immunization** |  |  |  |  |
| Yes | 62 (42.5) | 91 (47.9) | 0.98 | 0.322 |
| No | 84 (57.5) | 99 (52.1) |  |  |
| **Time spent at immunization service** |  |  |  |  |
| <30 minutes | 82 (56.2) | 115 (60.5) | 8.01 | **0.018** |
| 30 minutes to 1 hour | 41 (28.1) | 63 (33.2) |  |  |
| > 1 hour | 23 (15.7) | 12 (6.3) |  |  |
| **Educated how to manage vaccines side effects** |  |  |  |  |
| Yes | 44 (30.1) | 44 (23.2) | 2.08 | 0.149 |
| No | 102 (69.9) | 146 (76.8) |  |  |
| **Being informed when to come for next vaccination** |  |  |  |  |
| Yes | 102 (69.9) | 153 (80.5) | 5.13 | **0.024** |
| No | 44 (30.1) | 37 (19.5) |  |  |

In addition, the health system related factors which were found to be statistically associated with child full immunization included time spent at immunization service [X^2^(P-value) =8.01(0.018)] and being informed when to come for next vaccination [X^2^(P-value) =5.13(0.024)].

When multivariate analysis was performed as shown in Table 6, it was noted that there was no association between all variables and child full immunization except place of delivery, which showed that children whose places of delivery was the health facility, were 2.12 times more likely to be fully immunized compared to children who were born at home [AOR (95%CI) p-value=2.12(1.14-3.94)0.017].

**Table 6:**
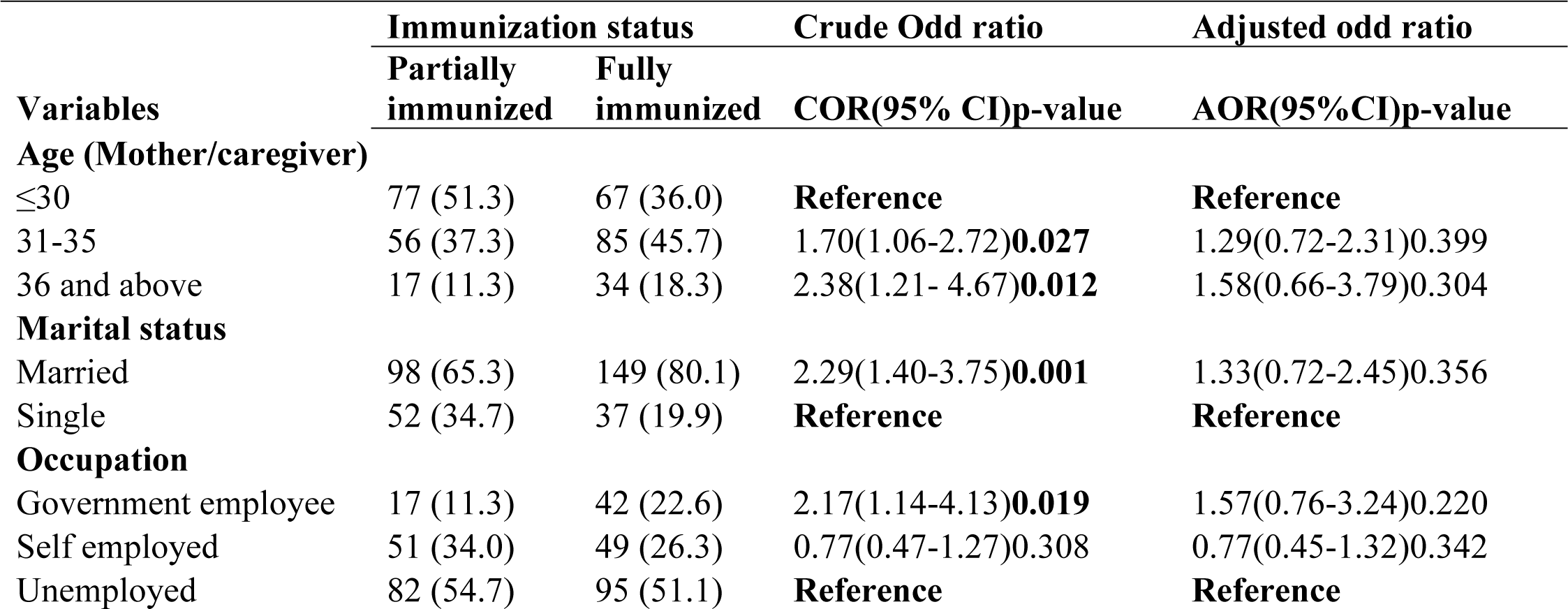
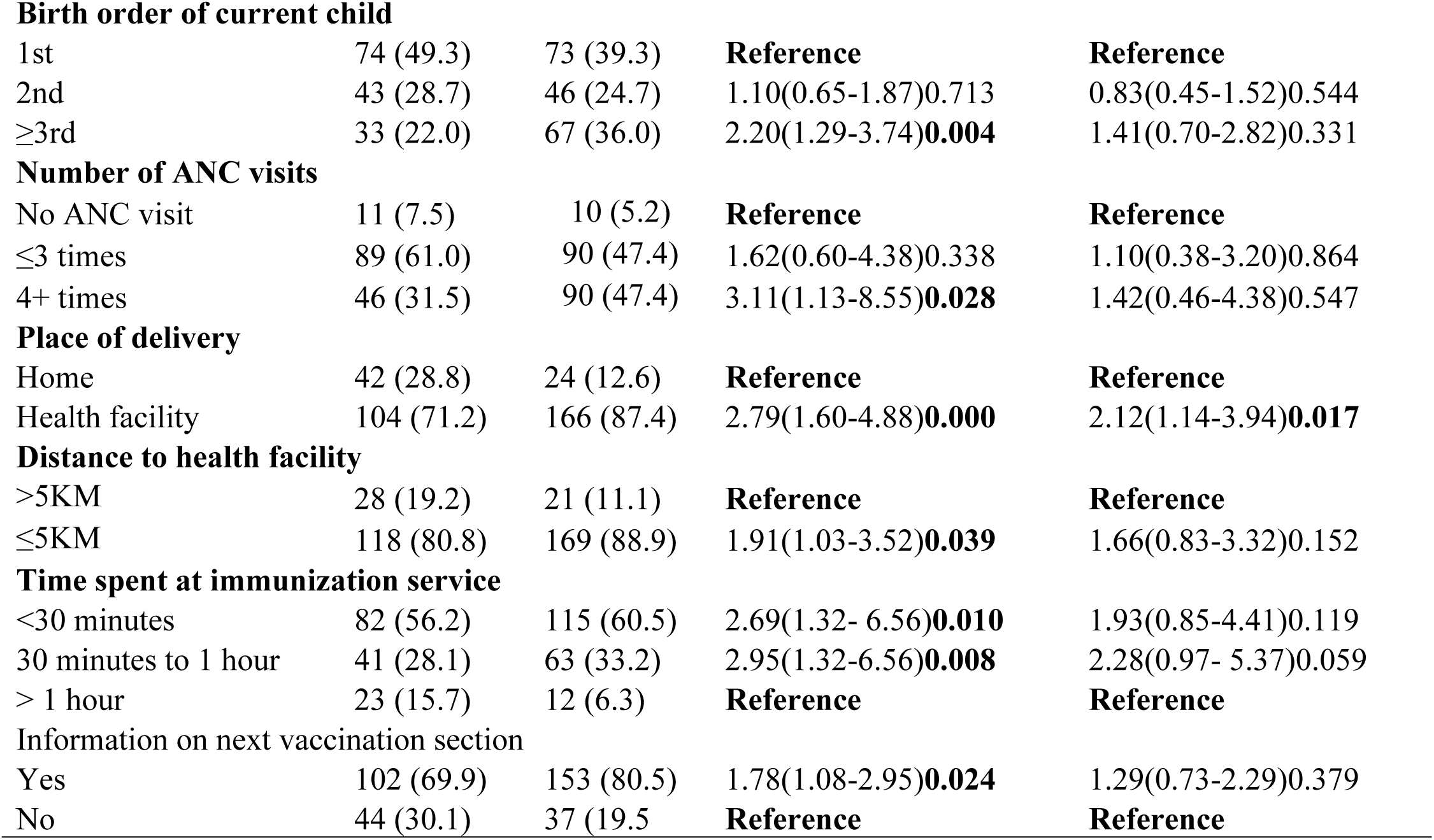
Predictors of immunization status among children under five years old in Nadowli-Kaleo District

| Variables | Immunization status |  | Crude Odd ratio | Adjusted odd ratio |
| --- | --- | --- | --- | --- |
|  | Partially immunized | Fully immunized | COR(95% CI)p-value | AOR(95%CI)p-value |
| **Age (Mother/caregiver)** |  |  |  |  |
| ≤30 | 77 (51.3) | 67 (36.0) | **Reference** | **Reference** |
| 31-35 | 56 (37.3) | 85 (45.7) | 1.70(1.06-2.72)**0.027** | 1.29(0.72-2.31)0.399 |
| 36 and above | 17 (11.3) | 34 (18.3) | 2.38(1.21- 4.67)**0.012** | 1.58(0.66-3.79)0.304 |
| **Marital status** |  |  |  |  |
| Married | 98 (65.3) | 149 (80.1) | 2.29(1.40-3.75)**0.001** | 1.33(0.72-2.45)0.356 |
| Single | 52 (34.7) | 37 (19.9) | **Reference** | **Reference** |
| **Occupation** |  |  |  |  |
| Government employee | 17 (11.3) | 42 (22.6) | 2.17(1.14-4.13)**0.019** | 1.57(0.76-3.24)0.220 |
| Self employed | 51 (34.0) | 49 (26.3) | 0.77(0.47-1.27)0.308 | 0.77(0.45-1.32)0.342 |
| Unemployed | 82 (54.7) | 95 (51.1) | **Reference** | **Reference** |
| **Birth order of current child** |  |  |  |  |
| 1st | 74 (49.3) | 73 (39.3) | **Reference** | **Reference** |
| 2nd | 43 (28.7) | 46 (24.7) | 1.10(0.65-1.87)0.713 | 0.83(0.45-1.52)0.544 |
| ≥3rd | 33 (22.0) | 67 (36.0) | 2.20(1.29-3.74)**0.004** | 1.41(0.70-2.82)0.331 |
| **Number of ANC visits** |  |  |  |  |
| No ANC visit | 11 (7.5) | 10 (5.2) | **Reference** | **Reference** |
| ≤3 times | 89 (61.0) | 90 (47.4) | 1.62(0.60-4.38)0.338 | 1.10(0.38-3.20)0.864 |
| 4+ times | 46 (31.5) | 90 (47.4) | 3.11(1.13-8.55)**0.028** | 1.42(0.46-4.38)0.547 |
| **Place of delivery** |  |  |  |  |
| Home | 42 (28.8) | 24 (12.6) | **Reference** | **Reference** |
| Health facility | 104 (71.2) | 166 (87.4) | 2.79(1.60-4.88)**0.000** | 2.12(1.14-3.94)**0.017** |
| **Distance to health facility** |  |  |  |  |
| >5KM | 28 (19.2) | 21 (11.1) | **Reference** | **Reference** |
| ≤5KM | 118 (80.8) | 169 (88.9) | 1.91(1.03-3.52)**0.039** | 1.66(0.83-3.32)0.152 |
| **Time spent at immunization service** |  |  |  |  |
| <30 minutes | 82 (56.2) | 115 (60.5) | 2.69(1.32- 6.56)**0.010** | 1.93(0.85-4.41)0.119 |
| 30 minutes to 1 hour | 41 (28.1) | 63 (33.2) | 2.95(1.32-6.56)**0.008** | 2.28(0.97- 5.37)0.059 |
| > 1 hour | 23 (15.7) | 12 (6.3) | **Reference** | **Reference** |
| **Information on next vaccination section** |  |  |  |  |
| Yes | 102 (69.9) | 153 (80.5) | 1.78(1.08-2.95)**0.024** | 1.29(0.73-2.29)0.379 |
| No | 44 (30.1) | 37 (19.5) | **Reference** | **Reference** |

## Discussion

Childhood immunization is crucial in stopping numerous illnesses and preventing an estimated 2 to 3 million deaths per year globally [27]. However, despite the effectiveness of vaccines, vaccine-preventable diseases still cause around 2.5 million childhood deaths annually, with roughly 1.5 million of those deaths occurring in developing countries among children under the age of 5 [28]. The main reason for conducting this study was to identify the factors that are associated with low childhood immunization coverage among mothers or caregivers. In this study, many factors have been found to influence childhood immunization uptake and are categorized below.

This our current study has found that the majority (55.4%) of respondents in this study were immunized fully. This present study’s coverage is lower, compared to the Ghana national target for all antigens of 95% coverage [14]. This is an indication that there are still high dropout rates of some antigens and could be indicative of poor performance and low access to immunization services. This finding did not agree with a lower figure that was reported in Ethiopia which showed that 77.4% of the children were fully immunized, and 15.5% were partially immunized [6]. However, a lower proportion of the overall full immunization coverage of 38.3% was reported in another study conducted in Ethiopia [29]. A similar finding was disclosed in a cluster survey that was carried out in the Techiman Municipality. The survey indicated that 89.5% of the children had received complete vaccinations, 9.5% had received some of the required vaccinations, and 1.0% had not been vaccinated at all [30].

According to the current study, age, marital status, occupation of the mother or caregiver, and birth order of the child being immunized were socio-demographic elements that showed a significant association with child complete immunization in the study area. This may be because mothers aged 19 and above may have become more aware of the significance of vaccination, leading to a decrease in the number of unvaccinated children. Additionally, the detrimental effects that result from not being immunized may have contributed to this positive trend [31]. Moreover, a likely reason regarding maternal occupation would be that mothers who might be engaged in one occupation or the other would not have to depend on their partners for transportation costs to take their children for immunization. The educational level of a mother could be improved by being married, as married mothers access to education may be better than single mothers with different responsibilities and may prioritize their children’s needs over their education [32]. If both partners collaborate to improve their child’s health, the supportive role of the partner may also boost the mother’s knowledge. This is similar to what was reported in Nigeria that maternal age, marital status, and occupation were some of the factors which all directly affected commencement, continuation, and completion of immunization [33]. Another similar finding was reported in the Assin North Municipality, Ghana, that the coverage of immunization was influenced by various maternal factors, such as the age, profession, educational background, and awareness regarding immunization and diseases that can be prevented through vaccination [21]. This finding of the current study, however, contradicted with what was reported in Burkina Faso, that the status of vaccination was considerably linked with factors such as place of dwelling, year of visit, ethnic background, and level of education of the mother [20].

Our study found that approximately 47.4% of study participants had their children fully immunized. Attending at least one ANC had significant association with complete vaccination. The results also showed that the majority (87.4%) of those who delivered in health facilities had their children fully immunized. This suggests that educating mothers about the benefits of immunizing their children early might have resulted in this. Furthermore, mothers who attended health care facilities regularly during pregnancy may have received childhood immunization counseling, which encouraged them to accept and prioritize the importance of timely childhood immunization. This finding further indicates that place of birth has a statistically significant association with full vaccination of children, and that children born in health care facilities are 2.12 times more likely to be vaccinated than those born at home. This is partly in support of the finding from a cross-sectional study reported in Kenya where the factors influencing low immunization coverage were found to be family size, place of birth, caregiver literacy, and their awareness, as well as whether you live a nomadic lifestyle [5]. A child born in a health facility is five times more likely to be vaccinated than a vaccine delivered to her home [5]. Moreover, in this present study, mothers who walked 5 kilometres or less to the nearest health facility had most of their children fully immunized (88.9%). Additionally, the distance to a health facility was significantly associated with child full immunization. This could be due to the fact that mothers who may be willing to get their children immunized and may be far from the reach of any health facility during the particular period of time and may not be able to afford transportation or walk to these outreach centres for services. This finding also agreed with the report of a study in Ethiopia, that distance to a health facility, delivery place, and follow-up during ANC were found to be significantly associated with children full immunization status [6].

In addition, it was found in this current study that the majority (52.1%) of the mothers who were never screamed at by health worker (s) when they were late during immunization had their children fully immunized. This may enhance or encourage mothers to take their children for vaccination. This finding further explained the revelation in a descriptive cross-sectional study conducted in Kenya, that the factor that hampers the process of regular immunization was attributed to the negative attitude of certain healthcare workers [17]. However, being screamed at by a health worker was not statistically associated with child full immunization status in this study. Moreover, time spent during immunization services was found to be statistically associated (p-value= 0.018) with child full immunization with a greater proportion of 60.5% of the mothers who spent less than 30 minutes at immunization had their children fully immunized. This supported the findings reported in rural areas of Nigeria that parent objections, disagreement, or concerns about immunization safety (38.8%), long distance walking (17.5%), and long waiting time at the health facility (15.2%) were the most common reasons for partial immunization [34].

Moreover, although not statistically associated, a greater proportion (76.8%) of mothers who were educated on how to manage vaccine side effects had their children fully immunized. Furthermore, being informed when to come for the next vaccination was also found to be significantly associated with child full immunization with the majority (80.5%) of the mothers who were informed about their next CWC visit had their children fully immunized. These findings of the present study were in consonance with what was reported in a community based cross-sectional study conducted in the slum areas of Mumbai, India, which reported that poor awareness and fear of adverse events, were some of the major reasons to not get immunized [19].

In conclusion, the immunization status of children in the Nadowli-Kaleo District was just a little above average (55.4%), hence, still below target. This study found that the factors which were significantly associated with child full immunization were age of mother or caregiver, marital status, occupation, and birth order of the current child. The results further showed that number of ANC visits, place of delivery, and distance to health facility were factors which had statistically significance association with child full immunization and hindered immunization. Children who were delivered in a healthcare facility had about 2.1 times greater chance of receiving full immunization as compared to those who were born in homes. The study also identified health system-related factors that had a significant correlation with complete immunization of children included time spent at the immunization service and being informed when to come for the next vaccination.

We therefore recommend that midwives and community health nurses should educate pregnant mothers to have skilled delivery, particularly at health facilities to ensure there is improved immunization coverage. In addition, the Nadowli-Kaleo District Health Directorate together with the Ghana Expanded Programme on immunization should conduct an immunization campaign frequently, focusing on all required vaccines and team up with sensitization of parents about the importance of completing the immunization schedule for children.

## Data Availability

Data for this study have been submitted to the Editor and can be obtained through a formal request from the Journal or through the corresponding author at

## Acknowledgement

Many thanks go to the District Director of Health Services, Nadowli-Kaleo District, the In-charges of all facilities who assisted me in obtaining the data and all my study participants (mothers/caregivers) for their cooperation and willingness to take part in the study, as well as my data collection assistants for helping me administer the questionnaires successfully.

## Supporting information

S1 Fig 1: Immunization status of children

S2 Cleaned Data used for analysis

S3 Stata Dofile for Data used for analysis

## References

1. Pan American Health Organization (PAHO), World Health Organization (WHO). Immunization [Internet]. 2022 [cited 2022 Nov 1]. Available from: https://www.paho.org/en/topics/immunization#:∼:text=Immunization%20is%20the%20process%20whereby,against%20subsequent%20infection%20or%20disease.

2. Center for Disease Control and Prevention (CDC). Understanding How Vaccines Work [Internet]. 2021 [cited 2022 Nov 1]. p. 3–5. Available from: https://www.cdc.gov/vaccines/hcp/conversations/understanding-vacc-work.html#print

3. WHO. Immunization coverage cluster survey, reference manual. 2005.

4. Rodrigues CMC, Plotkin SA, Foundation MG. Impact of Vaccines ; Health, Economic and Social Perspectives. 2020;11(July).

5. Kiptoo E, Esilaba M, Kobia G, Ngure R. Factors Influencing Low Immunization Coverage Among Children Between 12 - 23 Months in East Pokot, Baringo Country, Kenya. Int J Vaccines Vaccin. 2015;1(2).

6. Girmay A, Dadi AF. Full Immunization Coverage and Associated Factors among Children Aged 12-23 Months in a Hard-to-Reach Areas of Ethiopia. Int J Pediatr [Internet]. 2019;2019:1–8. Available from: 10.1155/2019/1924941

7. WHO. Immunization coverage: Facts sheet [Internet]. 2019 [cited 2020 May 15]. Available from: https://www.who.int/news-room/fact-sheets/detail/immunization-coverage

8. Bangura JB, Xiao S, Qiu D, Ouyang F, Chen L. Barriers to childhood immunization in sub-Saharan Africa: A systematic review. BMC Public Health. 2020;20(1).

9. Kazungu JS, Adetifa IMO. Crude childhood vaccination coverage in West Africa: Trends and predictors of completeness [version 1; Referees: 1 approved, 3 approved with reservations]. Wellcome Open Res. 2017;2(0):1–18.

10. Almond D, Currie J. Human Capital Development before Age Five. In: Handbook of Labor Economics. 2011. p. 1315–486.

11. Ministry of Health - Ghana. Immunization Programme Comprehensive Multi-year Plan (2010--2014) [Internet]. Accra; 2014. Available from: http://bidinitiative.org/wp-content/files_mf/1405555264GhanaComprehensivemultiyearplanfor20102014YearUnknown.pdf

12. Zewdie A, Letebo M, Mekonnen T. Reasons for defaulting from childhood immunization program: A qualitative study from Hadiya zone, Southern Ethiopia. BMC Public Health [Internet]. 2016;16(1):1–9. Available from: 10.1186/s12889-016-3904-1

13. Konwea PE, David FA, Ogunsile SE. Determinants of compliance with child immunization among mothers of children under five years of age in Ekiti State, Nigeria. J Heal Res. 2018;32(3):229–36.

14. Yawson AE, Bonsu G, Senaya LK, Yawson AO, Eleeza JB, Awoonor-Williams JK, et al. Regional disparities in immunization services in Ghana through a bottleneck analysis approach: Implications for sustaining national gains in immunization. Arch Public Heal [Internet]. 2017;75(1):1–10. Available from: 10.1186/s13690-017-0179-7

15. WHO/UNICEF. Improving Routine Immunization Service Delivery to Urban Poor in Ghana : Results of Situational Analysis AREAS SAMPLED. 2017;(July).

16. DHIMS2. Pent 3 coverage. 2019.

17. Njeru SK, Kagoiyo WS, Butto D. Barriers To Uptake of Childhood Routine Immunization In Nyeri County, Kenya. IOSR J Nurs Heal Sci. 2017;6(2):79–85.

18. Noh J, Kim Y, Akram N, Yoo K, Park J, Cheon J, et al. Factors affecting complete and timely childhood immunization coverage in Sindh, Pakistan; A secondary analysis of cross-sectional survey data. PLoS One [Internet]. 2018;13(10):1–15. Available from: 10.1371/journal.pone.0206766

19. Singh S, Sahu D, Agrawal A, Vashi MD. Barriers and opportunities for improving childhood immunization coverage in slums: A qualitative study. Prev Med Reports [Internet]. 2019;14. Available from: 10.1016/j.pmedr.2019.100858

20. Kagoné M, Yé M, Nébié E, Sie A, Schoeps A, Muller O, et al. Vaccination coverage and factors associated with adherence to the vaccination schedule in young children of a rural area in Burkina Faso. Glob Health Action [Internet]. 2017;10(1). Available from: 10.1080/16549716.2017.1399749

21. Ofosu SK. Factors Contributing To Immunisation Coverage in Assin North Municilpality [Internet]. University of Ghana; 2017. Available from: http://student.ucol.ac.nz/library/onlineresources/Documents/APA_Guide_2017.pdf

22. Ibrahim IS. Routine Infant Immunization ; Factors Influencing Noncompliance With the Schedule in the Tamale Metropolis. University for Development Studies; 2017.

23. Tadesse T, Getachew K, Assefa T, Ababu Y, Simireta T, Birhanu Z, et al. Factors and misperceptions of routine childhood immunization service uptake in ethiopia: Findings from a nationwide qualitative study. Pan Afr Med J. 2017;28:1–9.

24. GSS. 2010 Population and housing Census:District analytical report. Accra; 2014.

25. Cochran WF. SamplingTechniques: Chapter 5. In: Sampling Techniques [Internet]. third edit. New York: John Wiley and Sons; 1977. p. 10. Available from: https://scholar.google.com.tr/scholar?q=sampling+techniques&btnG=&hl=en&as_sdt=0,5#0

26. Nath DC, Patowari B. Estimation and Comparison of Immunization Coverage under Different Sampling Methods for Health Surveys. Int J Popul Res. 2014;2014(i):1–14.

27. United Nations Children’s Fund (UNICEF). Vaccination and Immunization Statistics - UNICEF DATA [Internet]. 2022 [cited 2023 May 4]. Available from: https://data.unicef.org/topic/child-health/immunization/

28. Aalemi AK, Shahpar K, Mubarak MY. Factors influencing vaccination coverage among children age 12-23 months in Afghanistan: Analysis of the 2015 Demographic and Health Survey. PLoS One [Internet]. 2020;15(8 August):1–16. Available from: 10.1371/journal.pone.0236955

29. Tamirat KS, Sisay MM. Full immunization coverage and its associated factors among children aged 12-23 months in Ethiopia: Further analysis from the 2016 Ethiopia demographic and health survey. BMC Public Health. 2019;19(1):1–7.

30. Adokiya MN, Baguune B, Ndago JA. Evaluation of immunization coverage and its associated factors among children 12-23 months of age in Techiman Municipality, Ghana, 2016. Arch Public Heal. 2017;75(1):1–10.

31. Galadima AN, Zulkefli NAM, Said SM, Ahmad N. Factors influencing childhood immunisation uptake in Africa: a systematic review. BMC Public Health. 2021;21(1):1–20.

32. Pepin JR, Sayer LC, Casper LM. Marital Status and Mothers’ Time Use: Childcare, Housework, Leisure, and Sleep. Demography. 2018;55(1):107–133.

33. Oleribe O, Kumar V, Awosika-Olumo A, Taylor-Robinson SD. Individual and socioeconomic factors associated with childhood immunization coverage in Nigeria. Pan Afr Med J. 2017;26:1–14.

34. Abdulraheem Is, Onajole AT, Jimoh AAG, Oladipo AR. Reasons for incomplete vaccination and factors for missed opportunities among rural Nigerian children. J Public Heal Epidemiol [Internet]. 2011;3(April):194–203. Available from: http://www.academicjournals.org/article/article1379427155_Abdulraheemetal.pdf

